# Rapid detection of SARS-CoV2 by Ambient Mass Spectrometry Techniques

**DOI:** 10.1101/2020.10.07.20207647

**Authors:** L. Ford, D. Simon, J. Balog, N. Jiwa, J. Higginson, E. Jones, S. Mason, V. Wu, E. Manoli, S. M. Stavrakaki, J. McKenzie, D. McGill, H. Koguna, J. Kinross, Z. Takats

## Abstract

Ambient Ionisation Mass Spectrometry techniques: Desorption Electrospray Ionisation (DESI) and Laser Desorption – Rapid Evaporative Ionisation Mass Spectrometry (LD-REIMS) were used to detect the SARS-CoV-2 in dry nasal swabs. 45 patients were studied from samples collected between April – June 2020 in a clinical feasibility study. Diagnostic accuracy was calculated as 86.7% and 84% for DESI and LD-REIMS respectively. Results can be acquired in seconds providing robust and quick analysis of COVID-19 status which could be carried out without the need for a centralised laboratory. This technology has the potential to provide an alternative to population testing and enable the track and trace objectives set by governments and curtail the effects of a second surge in COVID-19 positive cases. In contrast to current PCR testing, using this technique there is no requirement of specific reagents which can cause devastating delays upon breakdowns of supply chains, thus providing a promising alternative testing method.

---

Mass testing combined with effective, intensive contact tracing is essential if the global severe acute respiratory syndrome coronavirus 2 (SARS-CoV-2) pandemic is to be controlled prior to the delivery of a vaccine.^1–3^ Reverse transcriptase polymerase chain reaction (RT-PCR) testing for COVID19 is currently the gold standard diagnostic test; it extracts viral RNA from a nasopharyngeal swab, which is then amplified and measured by detection of a fluorescent tag. However, this analysis is slow (2 to >12 hours), costly and it has an inadequate sensitivity. Many governments failed to stock pile reagents prior to the pandemic, and those countries with centralised testing facilities have been unable to meet demand.^4^ In response to the pandemic, multiple diagnostic variations of Nulceic Acid Tests (NAT) methodologies have been rapidly developed as have protein tests based on the analysis of immunoglobulins or viral antigens.^5^ However, many of these suffer from similar challenges and RT-PCR and they have yet to be deployed at a population level. The United Kingdom’s government has therefore announced a testing Moon Shot programme to meet the mass testing challenge for COVID19.^6^

Mass spectrometry (MS) represents a scalable and viable alternative to RT-PCR and antigen analysis for mass testing. Matrix Assisted Laser Desorption Ionisation (MALDI) has completely revolutionised clinical microbiology^7^ and it has shown high diagnostic accuracy from nasal swabs when analysed using support vector machine learning algorithms.^8^ Metabolomics approaches using UPLC-MS to analyse sera have identified 204 metabolites which correlate to disease severity in COVID-19 patients.^9^ In fact, several MS based methodologies have been recently described which suggest that both metabonomic and proteomics have significant diagnostic applications.^10–12^ A global MS coalition has now been established to accelerate the development of diagnostic and prognostic biomarkers.^13^

One obstacle to the translation of MS based in vitro diagnostics is the time taken for sample preparation. Ambient mass spectrometry techniques such as Desorption Electrospray Ionisation (DESI) and Rapid Evaporative Ionisation Mass Spectrometry (REIMS) directly analyse samples (Figure 1). Desorption electrospray ionisation (DESI) utilises an electrospray emitter to create gas phase ions, ionic clusters and charged microdroplets which are directed at a candidate sample.^14^ Rapid evaporative ionisation mass spectrometry (REIMS) generates aerosolised droplets from the evaporation of sample by either Joule heating or laser irradiation.^15^ These two methods of mass spectrometry enable collection of m/z data representative of the surface of the biological sample in seconds. The speed, specificity and adaptability of these techniques enables potential for a fast and reliable point of care test for COVID-19. Another advantage of this technique is that the samples can be analysed directly from the dry swab, meaning that there is little to no sample preparation and risk of infection from sample preparation to the analysts.

**Figure 1.**
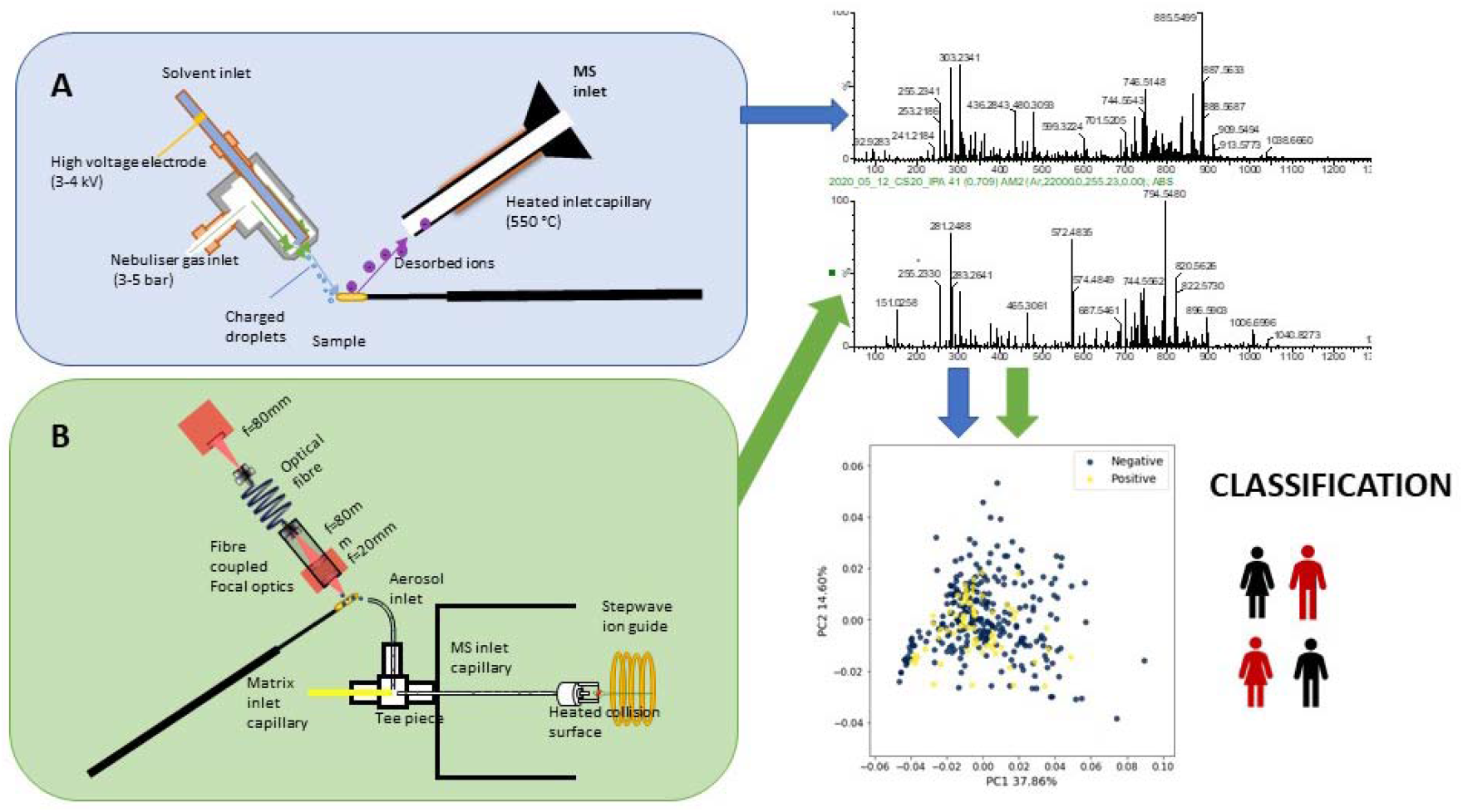
Schematic of MS workflow used in this project showing detailed schematic diagrams of A) DESI source where an electrospray emitter is used to produce gas phase solvent ions, ionic clusters and charged microdroplets.^14^ An electric potential is applied to the solvent in the electrospray emitter and this imparts charge on the surface molecules in the path of the solvent projectile and B) LD-REIMS source which uses a Q-switched Optical Parametric Oscillator laser emitting at 2940nm which rapidly heats the water within biological samples causing cells to explode, forming gas phase clusters of ions and droplets containing structural lipid information and metabolites specific to the biological matrix. Aspirated ions are then transported to the MS instrument for analysis. After this multivariate statistical analysis is performed and used to classify disease status.

These are mature technologies which have been widely deployed in multiple clinical domains such as surgery for real time tissue diagnostics^16–18^ and in microbiology^19^. This technology therefore has the potential to be implemented at the point of care or it could be leveraged for mass testing of COVID-19. In this clinical pilot study, the diagnostic accuracy of ambient mass spectrometry platforms were prospectively assessed for COVID-19 using nasopharyngeal swabs as a sampling methodology.

## Results

45 patients were prospectively recruited (median age 65 (25-87), 27 males and 18 females) (Table 1) as part of a feasibility study across Imperial College Healthcare Trust hospital sites in London between April 2020 and June 2020. The cohort contained 10 patients with active COVID-19 and a positive RT-PCR and 35 patients who were negative for COVID-19 on RT-PCR. Patients who had previously had a possible exposure to COVID-19 were included in this analysis, and of the 35 control patients, 21 had previously demonstrated a positive RT-PCR result, but none of these were within 7 days of the negative classification test used to build the diagnostic MS models. 4 patients in the control cohort had an active cough and 4 were experiencing shortness of breath. In the COVID-19 cohort, all subjects were in patients at the time of sampling, but none were ventilated or receiving ventilatory support.

**Table 1.** Demographics and characteristics of COVID 19 patients.

| Characteristics | Total (n = 45) | Positive (n = 10) | Negative (n = 35) |
| --- | --- | --- | --- |
| Age median (range) | 64 (25-87) | 62 (25-75) | 66 (32-87) |
| Male: Female | 27:18 | 4:6 | 23:12 |
| Previous exposure | 21* | 0 | 21* |
| Symptoms displayed |  |  |  |
| Cough | 4 | 0 | 4 |
| Tachypnoea | 2 | 2 | 0 |
| Dysphonia | 1 | 0 | 1 |
| Shortness of breath | 4 | 0 | 4 |
| Fever (>37.8<sup>0</sup>C) | 1 | 1 | 0 |
| Loss of smell and taste** | 0 | 0 | 0 |
| Radiological findings |  |  |  |
| CT or CXR with active COVID | 9 | 2 | 7 |
| CT or CXR with no active COVID | 19 | 5 | 14 |
| CT or CXR with residual signs of COVID/recovery | 7 | 0 | 7 |
| CT or CXR not performed | 10 | 3 | 7 |

\*Previous exposure= patients who had a positive RT-PCR test >7 days prior to the negative test used herein to classify patients as negative.
\*\*Loss of smell and taste was recognised in the UK 18<sup>th</sup> May 2020<sup>21</sup> and hence some swabs were taken prior to it being recognised, and therefore may have gone unrecorded by clinical staff.

RT-PCR was chosen as the gold standard method for distinguishing positive patients with active COVID19 from negative patients^20^ and it is part of routine clinical care carried out within Imperial College Healthcare Trusts. Patients who tested negative were followed up to observe if they developed COVID-19. One patient within the negative control cohort tested positive for COVID-19 within a week from the test swab. Patients underwent imaging at the discretion of the clinical team, and the radiological findings were compared to their corresponding PCR findings (Table 1). It was observed that 30% of those patients who yielded negative PCR and who underwent radiological investigation displayed positive radiological results suggestive of COVID-19.

It was hypothesised that dry nasal swabs would contain biological information on the inflammatory status of the respiratory tract of the host along with any viral or bacterial species present.

### Ambient Ionisation Mass Spectrometry Analysis

Due to precautions of transmission in the laboratory for the machine operators, swab samples were deactivated prior to analysis due to possible aerosolization of the virus in the analysis by ambient MS techniques. Deactivation is achieved by oven heating at 60°C for 30 mins.^22^ Figure 2 displays the effect of deactivation of the swabs on sample quality. From the spectra displayed, an increase in the ratio of biological material (in the region m/z 600-1000) to swab peaks is observed upon deactivation, possibly due to the swab being dried upon deactivation and therefore lower water content of the biological material being bombarded with the sprayer. It is important to note that the most abundant peaks in the spectra remain the same before and after deactivation and that there is no observable loss of abundant biological peaks. This experiment was achieved by the operator using their own swabs in an otherwise empty lab so therefore not contaminating anyone else with aerosolised particles from insufficiently deactivated samples.

**Figure 2.**
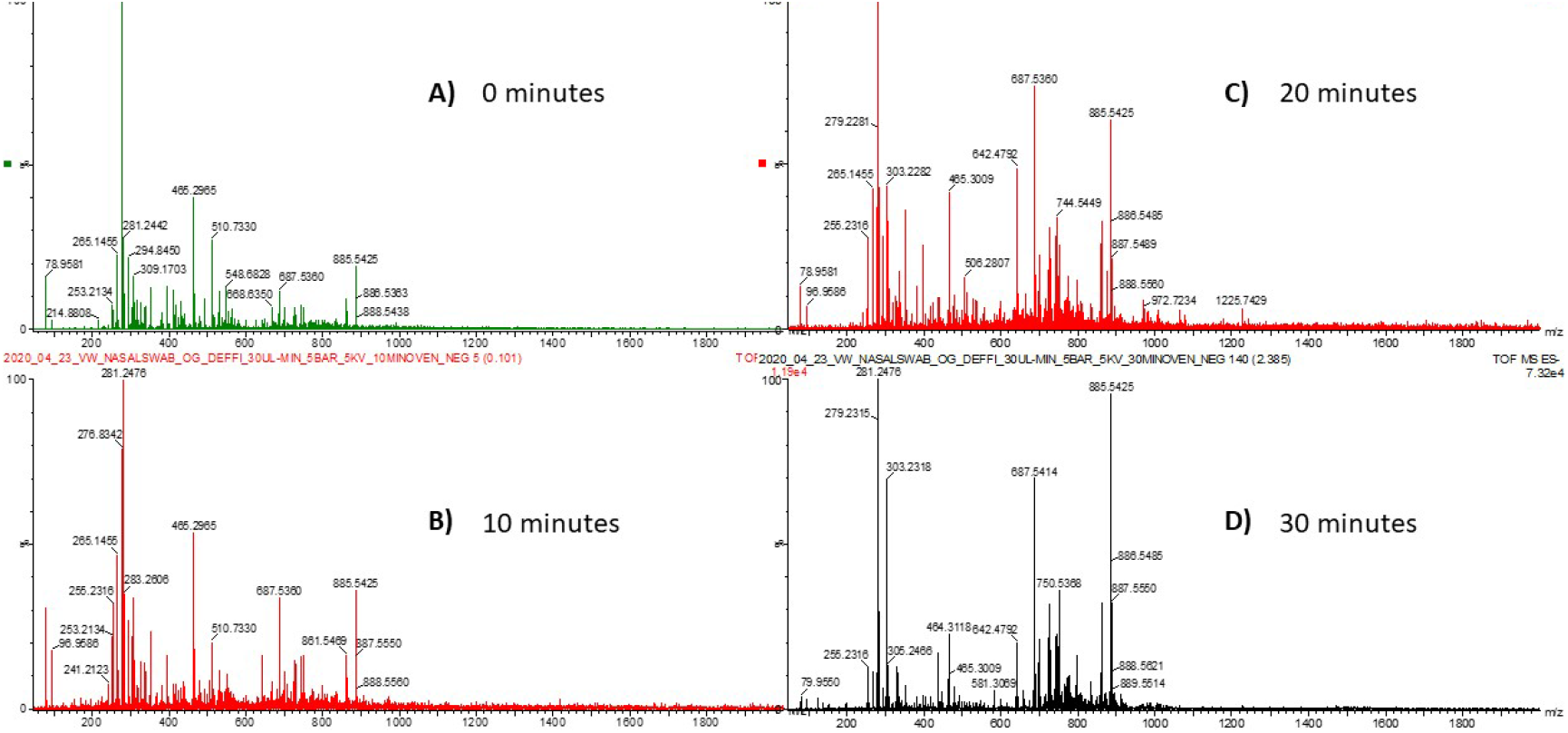
Effect of deactivation on DESI COVID-19 analysis after baking in the oven for A) 0 minutes, B) 10 minutes, C) 20 minutes and D) 30 minutes.

Once deactivated swab samples were analysed in sequence by DESI and LD-REIMS (Figure 3) so that a precise analysis of diagnostic accuracy could be performed. LD-REIMS is by its nature destructive, and therefore samples were first analysed by DESI. All samples were processed and analysed immediately after collection in order to keep the workflow as similar as possible to the point of care instrumentation that this work is demonstrating proof of concept for.

**Figure 3.**
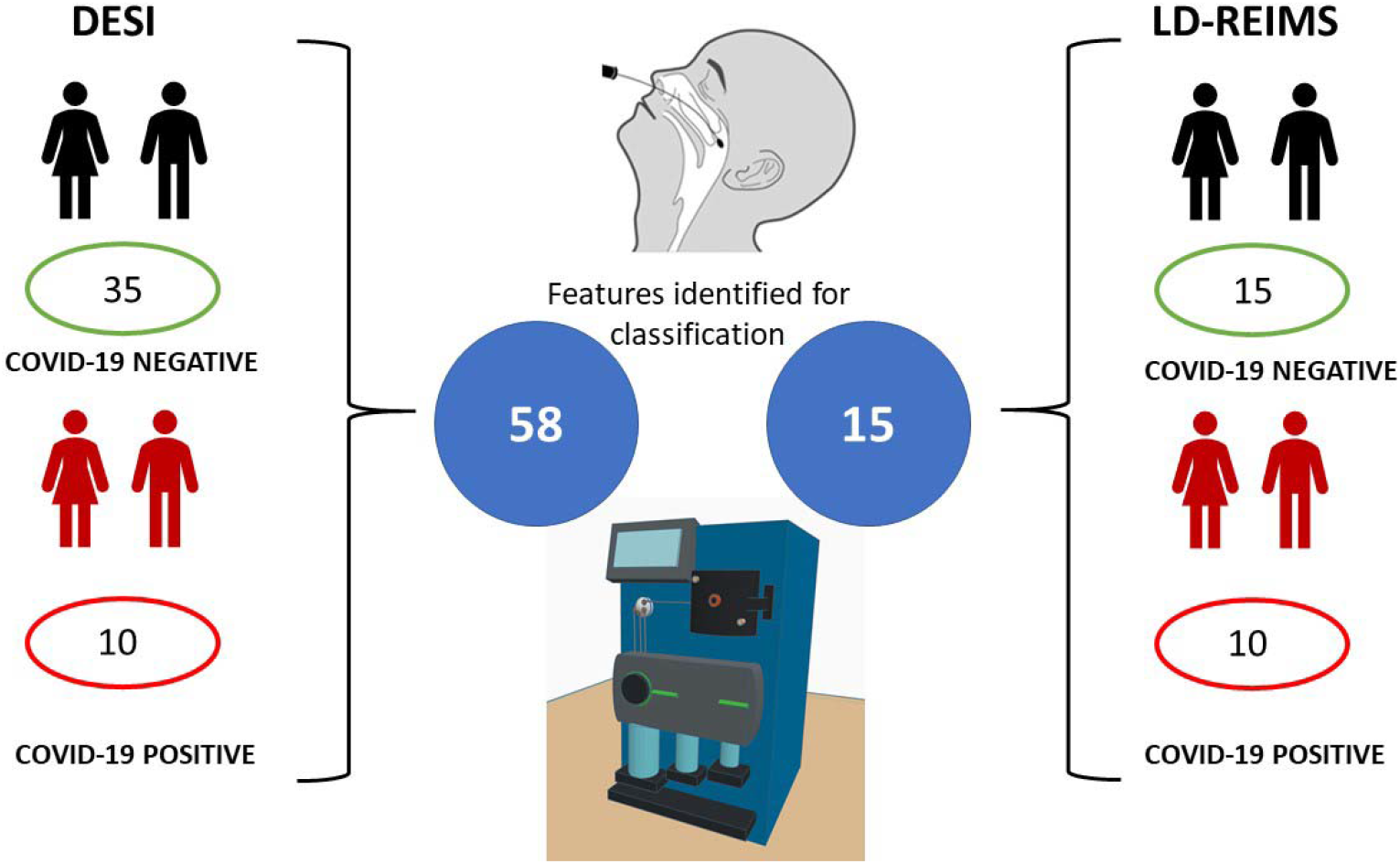
Schematic of Ambient Mass Spectrometry analysis pipeline for feature extraction, classification and diagnosis of COVID-19 from dry nasal swabs showing the demographics of patients recruited for each technology.

DESI was run directly on the deactivated dry swabs without any need for extraction or preparation. The data acquisition files were collected at 1 scan per second and collected in seconds, depending on operator and the quality of biological sample on the swabs.

DESI data was extracted using Waters Corporation software AMX and lockmass corrected to an internal peak of PI(38:4) which was present in all samples in high abundance. Total ion content was normalised, and the data was binned to m/z 0.1. Features were then selected using linear support vector classification (SVC) – resulting in 58 features, which were used for classification and cross validation. The features that contributed to this peak list are summarised in Supplementary Data Table 1. Figure 4 displays box plots of some of the DESI spectral features that drive classification between the positive or negative diagnosis classifications.

**Figure 4.**
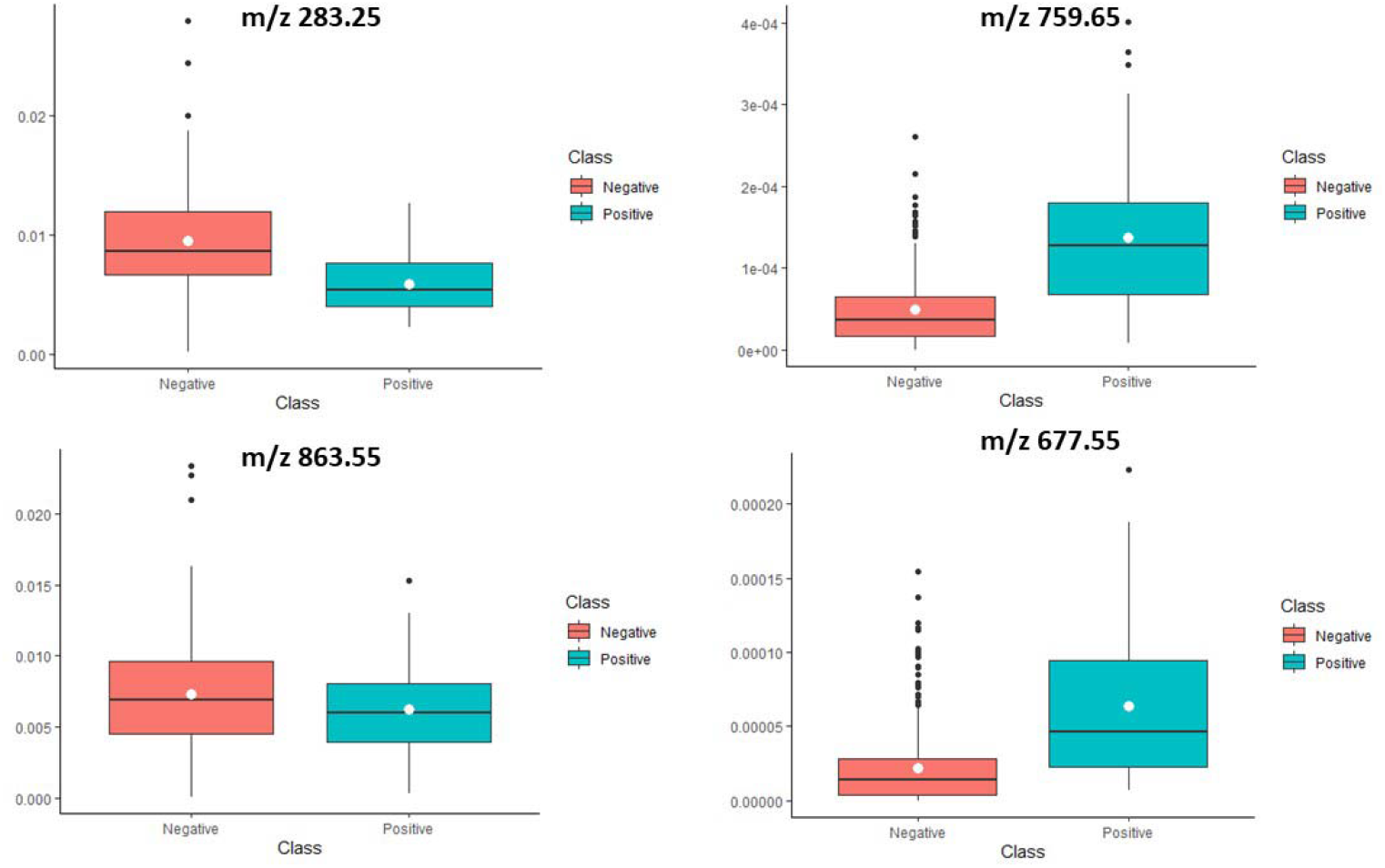
Box plots displaying intensity of lipid features which drive classification between positive and negative diagnosis in DESI analysis. The box represents the interquartile range with the mean intensity displayed as the white dot, the whiskers represent the range of data points excluding outliers, which were defined as more than 1.5 x the interquartile range from the mean.

DESI analysis showed high diagnostic accuracy when using the peak list of 58 features to classify patients using the SVC to classify patients. Using this supervised data analysis technique 39 patient samples were classified correctly out of 45 patients (Table 3, Supplementary Information). This gives a diagnostic accuracy of 86.7% which was calculated from leave one patient out cross validation. Given the sample cohort is small, this diagnostic accuracy was deemed to be acceptable and the model is expected to achieve higher diagnostic accuracy with increased number of samples. Tentative identification of the features in the peak list driving classification were performed by looking at the original lockmass corrected mass spectra to identify the accurate mass of the peak within the mass bin. This mass was then used to match to known species in databases such as lipidmaps and METLIN (Table 1, Supplementary information).

LD-REIMS was also used to classify a smaller cohort of samples; a smaller set of samples were used herein due to the availability of samples after COVID-19 risk assessments were completed and laser safety was allowed with reduced capacity of workers in laboratories. The data was treated in the same way as the DESI data, but the lockmass used was the internal lipid of palmitic acid. This is due to the different methods of excitation and ionisation of liberated ions between DESI and LD-REIMS, there is a different spectral pattern between the two techniques and hence the most abundant peaks between the two different techniques differ significantly, therefore different lockmass peaks were selected for the two different ambient mass spectrometry techniques. Due to the smaller cohort of samples subjected to this analysis, the peak list extracted from the support vector machine learning algorithm totalled 15 peaks.

LD-REIMS also showed high diagnostic accuracy supervised support vector machine learning classification. The technique correctly classified 21 out of 25 total patients, which results in a diagnostic accuracy of 84%. As with the DESI data, patient’s disease status is correlated to the patients most recent PCR test result. Figure 4 displays the unsupervised principle component analysis (PCA) along with the loadings plots from PC1, PC2 and PC3. DESI and LD-REIMS displayed similar variance in the data in PC1 of 37.86% and 34.01% respectively. PCA clustering was used to reduce the dimensions in the dataset for higher quality supervised classification techniques which calculate the relationship between the variables and separate based on factors that drive classification between classes.

### Spectral Content

Both DESI and LD-REIMS display different spectral patterns which can be observed in Figure 6. This is due to the differing mechanisms of aerosol formation and ionisation in the two different techniques (Figure 1). In DESI analysis, an electrospray emitter is used to produce gas phase solvent ions, ionic clusters and charged microdroplets.^14^ An electric potential is applied to the solvent in the electrospray emitter and this imparts charge on the surface molecules in the path of the solvent projectile. If the surface group is given enough momentum it will leave the surface, be transported through the air at atmospheric pressure and reach the atmospheric interface of the mass spectrometer. Unlike usual DESI applications in mass spectrometry imaging (MSI) where a very fine sprayer is required for better resolution, herein settings were optimised to spray a large surface area very quickly.

**Figure 5.**
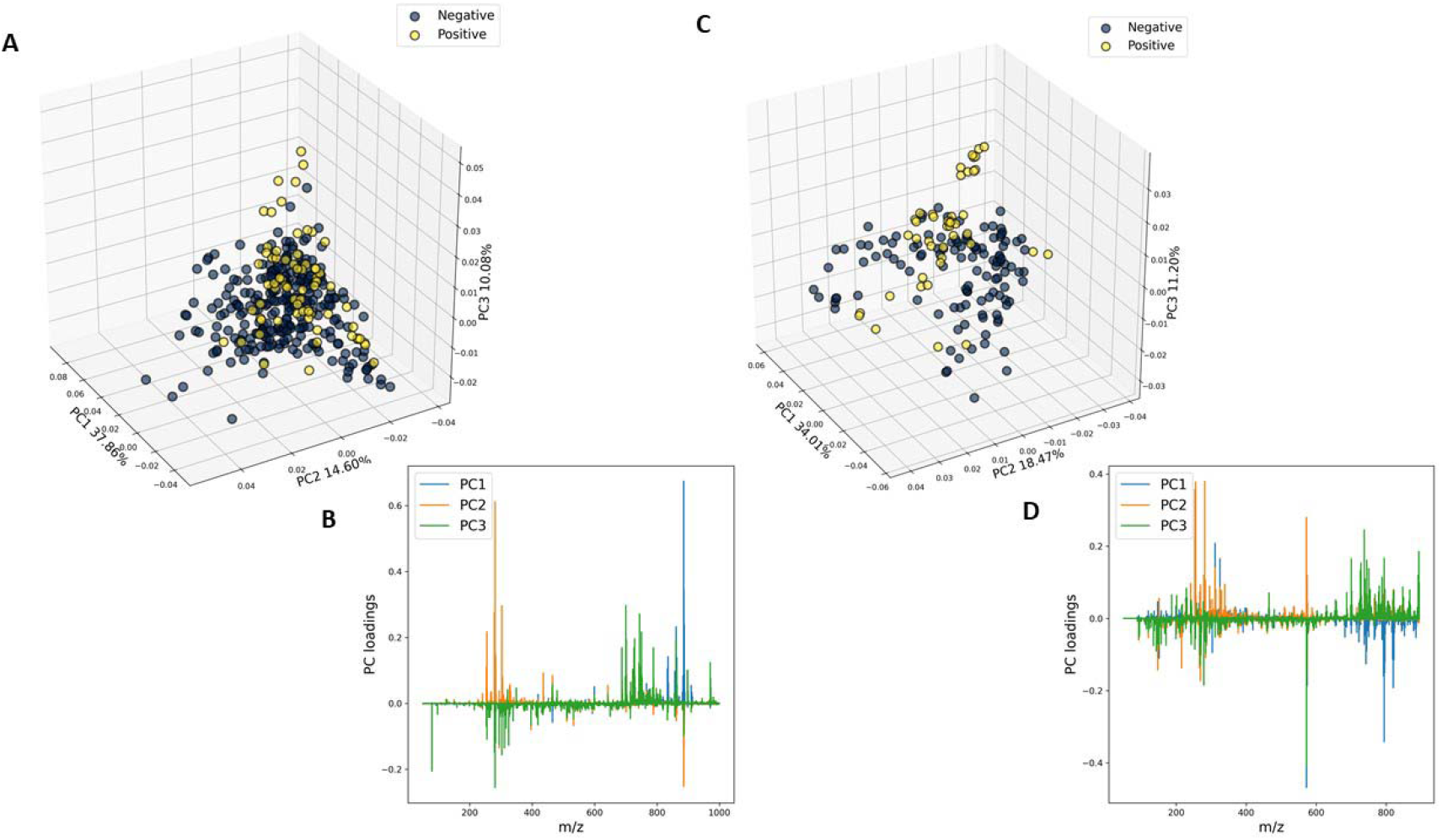
Data visualisation of ambient mass spectrometry data A) PCA of DESI data, B) loadings plots of DESI data, C) PCA of LD-REIMS data, D) loadings plots of LD-REIMS data.

**Figure 6.**
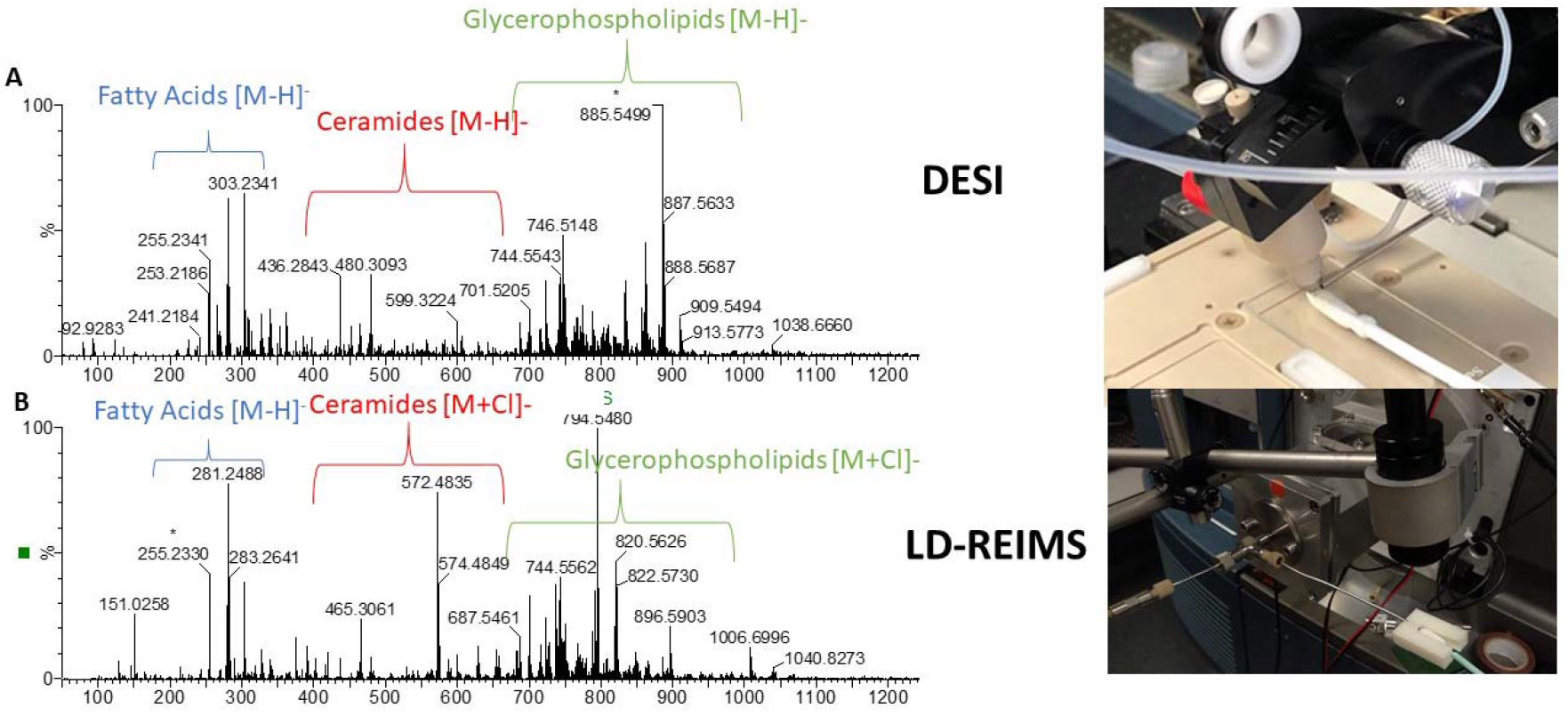
Sample spectra from nasopharyngeal COVID19 negative swab samples A) DESI and B) LD-REIMS. Displaying the differing spectral pattern between DESI and LD-REIMS.

LD-REIMS uses a Q-switched Optical Parametric Oscillator laser emitting at 2940nm which rapidly heats the water within biological samples causing cells to explode, forming gas phase clusters of ions and droplets containing structural lipid information and metabolites specific to the biological matrix. These aspirated metabolites are then transferred to the interface of the mass spectrometer where they are mixed with a matrix solvent, herein 2-isopropanol was used. The droplets which are absorbed into the matrix are then carried at high speed into the vacuum of the MS and encounter a heated collision surface maintained at 1200 K. Upon collision on the heated surface the droplets rapidly evaporate, and the ions are de-clustered allowing measurement of single ions.^23^

Due to the nature of mass spectrometry profiling, contrary to other methods such as optical spectroscopy, the peaks observed are due to individual masses of particular lipids. Therefore, the mass spectroscopy profile can be used as a fingerprint of disease status and has biological meaning. The increase or decrease of abundance of these lipids can be used to explain some of the biological pathways associated with disease status and the observations made in this analysis.

This analysis identified a decrease in abundance of free fatty acids and Phosphatidylinositol (PI) lipid species measured from swabs with positive COVID-19 status. This is in common with findings from other metabolomic studies focusing in COVID-19 disease states which saw general decreases in phospholipid and free fatty acid abundance in COVID-19 positive cases in blood plasma.^9^ Phospholipase enzymes, which cleave phospholipids into lysophospholipids and free fatty acids, have been linked to COVID-19 viral replication efficiency and immune response.^24^ These phospholipases are linked to strong immune response due to their role in production of eicosanoids from free fatty acids, in particular arachidonic acid (20:4) and hence this observation would be expected for infected patients. Platelet degranulation pathways associated with inflammation also cause significant changes in lipid abundance; and could be one of the major biochemical factors in the observed lipid species specific to COVID-19 diagnosis by ambient mass spectrometry techniques. The levels of phosphatylethanolamine (PE) and phosphatidylserine (PS) have been shown to be distributed asymmetrically in platelet bilayers. Activated externalisation of PE and PS phospholipids in platelet micro vesicles (MV) could also be the reason for their increased exposure in positive COVID-19 nasal fluid swabs.^25^ The increased abundance of these phospholipids in patients with positive COVID-19 diagnosis could be due to the formation of MV for viral replication in the cells. Increased coagulation is a common clinical observation in patients with COVID-19 and the relationship between platelet formation in the inflammatory response and the observed lipidomic species could be driving classification herein.^26^ Most omics studies performed so far on COVID-19 infected fluids have been focused on blood plasma samples^9,27^ and using different ionisation techniques which makes comparison difficult.

## Limitations

One of the major limitations of this study is the sample size, and hence, further work will look to significantly enlarge the sample cohort of patients for analysis and clinical validation of this study. Another limitation of developing COVID-19 omics-based diagnostics tools in general, is the lack of robust gold standard tests herein for training datasets in machine learning. PCR is used in this study, but the accuracy of this test is problematic; in this study alone 7 out of 35 patients that tested negative for COVID-19 displayed radiological results consistent with COVID-19 infection. However, 6 out of 7 of these patients had been previously exposed to COVID-19 testing positive >7 days previously. This suggests that radiology results display changes in the lung caused by COVID-19 but is not reflective of the patient’s current viral load. Whilst this study displays a variety of severity in the patients that tested positive for COVID-19, there was no study on the viral load and its effects on this model.

Furthermore, this study has only addressed the effect of COVID-19 pathological status in clinical detection. In a larger dataset, further work will look to analyse the effect of confounding pathologies such as common colds or flu on the COVID-19 metabolomics approach used herein.

Another factor to be considered is the aerosol generation produced by this analysis technique. Due to the nature of the aspiration in both DESI and LD-REIMS, 95% of the aspirated aerosol generated described in this work is organic solvent and should therefore deactivate any live virus as part of the normal workflow. As an extra precaution herein, all samples were deactivated before analysis. Future work encompasses the development of an air-tight chamber which encloses the swab and virus particles within it for analysis. Further work will also test the use of solvents such as IPA or methanol to immerse the swab before analysis. This mirrors the LD-REIMS workflow, but further validation for DESI techniques will establish if these alternative and faster deactivation methods are feasible. This will remove the need for deactivation and enable high-throughput analysis in a matter of seconds per sample.

## Conclusion

The ambient ionisation mass spectrometry techniques demonstrate high diagnostic accuracy of 86.7% and 84% for DESI and LD-REIMS respectively which is expected to increase with a larger cohort of samples. DESI and LD-REIMS are therefore promising tools for the rapid diagnosis of COVID-19 infection status. The sampling method can be achieved in seconds without the need for sample preparation, which when compared to gold standard PCR testing, cuts analysis time significantly. One other advantage of this technique is that once geometry of the DESI sprayer is optimised and set up, then the samples can be collected and run by point of need staff with minimal training and future iterations of the instrument are expected to run completely automated. This technology provides a revolutionary approach to traditional microbiology testing which is easily scaled to deal with mass population testing required for control of the coronavirus pandemic. The implementation of this technology removes the risk of supply chain shortages causing delays because it does not rely on specific reagents and primers providing a promising alternative test to the traditional PCR approach.

## Methods

### Recruitment of patients

Participants were prospectively recruited to this feasibility study within a sub-collection of Imperial College Tissue Bank, which acts under the HTA license 12275 and research ethics committee approval 17/WA/0161. Patients or staff at three London hospitals within Imperial College NHS Healthcare Trust between 29^th^ April and 30^th^ June 2020 were eligible for inclusion if they were aged 18 or over; displayed symptoms of COVID-19 (a new continuous cough, shortness of breath, temperature above 37.8 C and anosmia); required admission to the trust for emergency surgery or healthcare staff exposed to COVID-19. All participants gave written and informed consent for inclusion in the study. Participants included in the study were pseudonymized into a CVS number and each swab was given a CS number; most patients provided two swab samples. Clinical meta-data collected included demographics, presence of symptoms consistent with COVID-19 and date of all PCR tests with the corresponding result.

### Sampling of Patients

Deep nasal secretion samples were collected from both nostrils, using a woven polyester swab (harmony P3685), by appropriately trained clinical research personnel wearing PPE in accordance with local procedures. Patients with nasal obstruction due to medical devices such as nasogastric tubes, or to pre-existing distorted anatomy, had samples collected from the unobstructed nostril only. The swab was gently inserted to the mid turbinate level and allowed to rest for at least two seconds to allow absorption of nasal secretions before withdrawal. Samples were then triple sealed in a container labelled as containing ‘UN 3373 biological substance category B’, before being transported to the laboratory for immediate deactivation.

### Deactivation

Samples were deactivated prior to analysis to reduce risk to analysts carrying out the study. Deactivation was done in a CL2 medical safety cabinet fitted with a Thermo Scientific 650 W High Capacity Dryer set to 60°C. A glass thermometer was also placed inside the oven to verify the temperature being read by the digital display. Once samples arrived in the laboratory they were placed in the oven and left for 30 minutes in order to deactivate any live virus.^22^

### DESI analysis

DESI stage was set up with the following optimised parameters. The DESI solvent used was a mixture of methanol/water at a ratio of 95:5. Flow rate was set to 30 µL/min, with the gas set at 5 bar (N_2_). Sprayer geometry was set to 2-3 mm distance from the inlet capillary and 1 mm distance from the surface of the swab sample and at a 75° impact angle. The nozzle diameter was 200 µm and the capillary to nozzle diameter was 200 µm. Data was collected at 1 scan per second in continuum mode on a Waters Xevo G2-XS QToF mass spectrometer. Data was collected in the range of m/z 50-2000 for each sample.

### LD-REIMS analysis

After DESI analysis Laser Desorption - Rapid Evaporative Ionisation Mass Spectrometry (REIMS) analysis was performed on the swabs. Fundamentally the technique requires the samples to be rapidly heated using a Q-switched Optical Parametric Oscillator laser emitting at 2940nm. This process evaporates the sample creating an aerosol rich in molecular information. The aerosol is transferred in front of the MS inlet capillary, where the droplets are mixed with a matrix solvent (2-isopropanol). The solvent mixed droplets enter the vacuum of the instrument where they impact on a heated (1200K) collision surface. This highly energetic collision liberates free ions from the droplets which is analysed by a Waters Corporation Xevo G2-XS QToF mass spectrometer. Prior to use the mass spectrometer was calibrated using sodium formate in negative electrospray ionisation mode, following the manufacture’s standard operating instructions. All LD-REIMS data was collected in negative ionisation in the mass range of m/z 50 – 1500.

### Statistical analysis

All acquired raw mass spectrometry files were imported into Abstract Model Builder (AMX) software (Waters Research Centre, Budapest, Hungary) for prepossessing. First, the data scans were combined into spectra, resulting in multiple spectra for each sample. The spectra were background subtracted and lockmass corrected to internal lipid and phospholipid species of m/z 255.233 and 885.5499 for LD-REIMS and DESI respectively. After lockmass correction, spectra were total ion content (TIC) normalised and binned to 0.1 Da. Principle component analysis was performed on the first 10 components (Figure 5). Both PCA and LDA identify linear combinations of variables within a dataset. PCA is an unsupervised technique that does not consider class separation of samples. LDA, however looks to identify linear relationships between variables taking into consideration class groupings and explicitly separates data according to these linear discriminants. PCA is initially used to reduce the dimensionality of the dataset and then supervised techniques are used to identify the relationship between the variables and build the models. The pre-processed data matrix was exported from AMX into Anaconda Spyder Python 3.7 and the sklearn support vector machine linear SVC algorithm was used for feature selection with L1 penalty norm and C=500 regularization parameter for classifications. Features were then input back into AMX for leave one patient out cross validation and calculation of the diagnostic accuracy. Box plots of features were created using ggplot2 package in R.

## Data Availability

Data for this manuscript is uploaded in supplementary information

## Author Contributions

LF contributed to writing the manuscript.

LF, JH, NJ, HK, SM, EM contributed to recruitment of patients and samples.

LF, DS, VW, EM, SS, JK, ZT were responsible for the experimental design and method development of MS techniques.

JK, ZT were responsible for the strategy and planning of the work.

SM wrote the ethics for the study.

## Competing interests

There are no competing interests declared by authors.

## Notes

### Competing Interest Statement

The authors have declared no competing interest.

### Funding Statement

This article is independent research funded by the National Institute for Health Research (NIHR) Imperial Biomedical Research Centre (BRC). The views expressed in this publication are those of the author(s) and not necessarily those of the NHS, the National Institute for Health Research or the Department of Health

### Author Declarations

The project has been approved by Imperial College Healthcare Trust Tissue Bank (ICHTB). ICHTB HTA licence: 12275 REC approval: 17/WA/0161. Address: Imperial College Healthcare Tissue Bank Department of Surgery and Cancer, Charing Cross Hospital Fulham Palace Road London W6 8RF Tel: 020 3311 7173

