## Supplementary Information for "Rapid detection of SARS-CoV2 by Ambient Mass Spectrometry Techniques"

**Supplementary information**

**Table 2.** DESI peak list with tentative annotations

| **Mass bin** | **Mass observed _DESI** | **True Mass** | **Delta** | **ppm error** | **Adduct** | **Annotation** | **Source** |
| --- | --- | --- | --- | --- | --- | --- | --- |
| 78.958 | 78.9591 | 78.9602 | 0.0011 | 13.93126 | [M-H]- | Phosphonic acid, ion(2-) | METLIN |
| 124.008 | 124.0075 | 124.0074 | -1E-04 | -0.8064 | [M-H]- | L-Taurine | METLIN |
| 199.17 | 199.1826 | 199.1704 | 0.0122 | -61.2503 | [M-H]- | Dodecanoic acid |  |
| 253.217 | 253.222 | 253.2173 | 0.0047 | -18.5608 | [M-H]- | FA(16:1) | lipid maps |
| 255.233 | 255.2342 | 255.233 | 0.0012 | -4.70156 | [M-H]- | FA(16:0) | lipid maps |
| 265.149 | 265.1488 | 265.1479 | 0.0009 | -3.39432 | [M-H]- | Lauryl sulfate | METLIN |
| 279.233 | 279.2339 | 279.233 | 0.0009 | -3.2231 | [M-H]- | FA(18:2) | lipid maps |
| 281.248 | 281.2486 | 281.2486 | 0 | 0 | [M-H]- | FA(18:1) | lipid maps |
| 283.263 | 283.2635 | 283.2643 | 0.0008 | 2.824226 | [M-H]- | FA(18:0) | lipid maps |
| 295.227 | 295.2284 | 295.2279 | -0.0005 | -1.6936 | [M-H]- | 13(R)-HODE | METLIN |
| 303.233 | 303.233 | 303.233 | 0 | 0 | [M-H]- | FA(20:4) | lipid maps |
| 452.256 | 452.2773 | 452.2783 | 0.001 | 2.211033 | [M-H]- | LPE(16:0) | lipid maps |
| 464.314 | 464.313 | 464.3147 | 0.0017 | 3.661323 | [M-H]- | PE(O-18:1) | lipid maps |
| 557.455 | 557.4576 | 557.4575 | -1E-04 | -0.17939 | [M-H]- | FAHFA(O-36:4) | lipid maps |
| 599.32 | 599.3189 | 599.3202 | 0.0013 | 2.169129 | [M-H]- | LPI(18:0) | lipid maps |
| 642.487 | 642.4866 | 642.4951 | 0.0085 | 13.22985 | [M-H]- | GlcCer(d30:1)) | lipid maps |
| 650.161 | 650.1566 | 650.1389 | -0.0177 | -27.2242 | [M-H]- | Hemin | METLIN |
| 673.232 | 673.2324 | 673.2398 | 0.0074 | 10.99175 | [M-H]- | Halistinol Sulfonic Acid F | lipid maps |
| 678.472 | 678.4694 | 678.4717 | 0.0023 | 3.389983 | [M+Cl]- | GlcCer(d30:1) | lipid maps |
| 687.545 | 687.545 | 687.5447 | -0.0003 | -0.43634 | [M-H]- | PE-Cer(d36:1) | lipid maps |
| 700.528 | 700.5281 | 700.5287 | 0.0006 | 0.856497 | [M-H]- | PE(O-34:2) | lipid maps |
| 701.519 | 701.5186 | 701.5127 | -0.0059 | -8.41033 | [M-H]- | PA(36:1) | lipid maps |
| 722.513 | 722.5132 | 722.513 | -0.0002 | -0.27681 | [M-H]- | PE(O-36:5) | lipid maps |
| 725.519 | 725.5127 | 725.5127 | 0 | 0 | [M-H]- | PA(38:3) | lipid maps |
| 726.544 | 726.545 | 726.5443 | -0.0007 | -0.96346 | [M-H]- | PE(O-36:3) | lipid maps |
| 740.522 | 740.5178 | 740.5236 | 0.0058 | 7.832357 | [M-H]- | PE(36:3) | lipid maps |
| 744.556 | 744.5568 | 744.5549 | -0.0019 | -2.55185 | [M-H]- | PE(36:1) | lipid maps |
| 749.525 | 749.5251 | 749.5338 | 0.0087 | 11.60735 | [M-H]- | PG(34:0) | lipid maps |
| 750.544 | 750.543 | 750.5443 | 0.0013 | 1.732079 | [M-H]- | PE(O-38:5) | lipid maps |
| 770.507 | 770.4921 | 770.4978 | 0.0057 | 7.397869 | [M-H]- | PS(35:3) | lipid maps |
| 770.574 | 770.5635 | 770.5705 | 0.007 | 9.084261 | [M-H]- | PE(38:2) | lipid maps |
| 775.547 | 775.547 | 775.5495 | 0.0025 | 3.223531 | [M-H]- | PG(36:1) | lipid maps |
| 788.544 | 788.5447 | 788.5447 | 0 | 0 | [M-H]- | PS(36:1) | lipid maps |
| 795.518 | 795.5012 | 795.5182 | 0.017 | 21.37018 | [M-H]- | PG(38:5) | lipid maps |
| 802.571 | 802.5737 | 802.5756 | 0.0019 | 2.367384 | [M-H]- | PE(O-42:7) | lipid maps |
| 834.522 | 834.5214 | 834.5291 | 0.0077 | 9.226845 | [M-H]- | PS(40:6) | lipid maps |
| 861.55 | 861.5511 | 861.5499 | -0.0012 | -1.39284 | [M-H]- | PI(36:2) | lipid maps |
| 863.564 | 863.5645 | 863.5655 | 0.001 | 1.157991 | [M-H]- | PI(36:1) | lipid maps |
| 865.572 | 865.5659 | 865.5812 | 0.0153 | 17.6763 | [M-H]- | PI(36:0) | lipid maps |
| 883.533 | 883.5321 | 883.5342 | 0.0021 | 2.376824 | [M-H]- | PI(38:5) | lipid maps |
| 886.553 | 886.5528 | 886.5604 | 0.0076 | 8.572529 | [M-H]- | PS(44:8) (isotope of 885 based on LCMS) | lipid maps |
| 889.572 | 889.58 | 889.5812 | 0.0012 | 1.348951 | [M-H]- | PI(38:2) | lipid maps |
| 970.719 | 970.7134 | 970.7200 | 0.0066 | 6.799123 | [M-H]- | LacCer(d42:2) | lipid maps |

**Table 3.** LD-REIMS peak list with tentative annotations.

| **Mass bin** | **Mass observed REIMS** | **True Mass** | **Delta** | **ppm error** | **Adduct** | **Annotation** | **Source** |
| --- | --- | --- | --- | --- | --- | --- | --- |
| 119.95 | 119.9465 | 119.947 | 0.0005 | 4.1685251 | [M+Cl]^-^ | Sodium Nitrate | METLIN |
| 124.05 | 124.0078 | 124.0074 | -0.0004 | -3.225604 | [M-H]- | L-Taurine | METLIN |
| 151.05 | 151.0257 | - | - | - | - | unknown |  |
| 255.25 | 255.233 | 255.233 | 0 | 0 | [M-H]- | FA(16:0) | lipidmaps |
| 265.15 | 265.1484 | 265.1479 | -0.0005 | -1.885736 | [M-H]- | Lauryl Sulfate | METLIN |
| 279.05 | 279.2328 | 279.233 | 0.0002 | 0.7162482 | [M-H]- | FA(18:2) | lipidmaps |
| 281.25 | 281.2487 | 281.2486 | -1E-04 | -0.355557 | [M-H]- | FA(18:1) | lipidmaps |
| 295.25 | 295.2275 | 295.2279 | 0.0004 | 1.3548873 | [M-H]- | 13(R)-HODE | METLIN |
| 303.25 | 303.2326 | 303.233 | 0.0004 | 1.3191194 | [M-H]- | FA(20:4) | lipidmaps |
| 325.15 | 325.1848 | - | - | - | - | unknown |  |
| 329.25 | 329.2445 | 329.2486 | 0.0041 | 12.452752 | [M-H]- | FA(22:5) | lipidmaps |
| 572.45 | 572.4152 | - | - | - | - | unknown |  |
| 603.45 | 603.452 | 603.4395 | -0.0125 | -20.71416 | [M-H]- | PA(O-30:0) | lipidmaps |
| 631.55 | 631.5005 | 631.4821 | -0.0184 | -29.13695 | [M-H]- | PE-Cer(32:1) | lipidmaps |
| 742.55 | 742.5336 | 742.5392 | 0.0056 | 7.5417463 | [M-H]- | PE(36:2) | lipidmaps |

**Table 4.** Patient details and classification for DESI data

| Patient | No. of spectra | No. of passes | No. of failures | No. of outliers | Correct Classification Rate (Exluding outliers) | Correct Classification Rate (Including outliers) | True Classification |
| --- | --- | --- | --- | --- | --- | --- | --- |
| CVS001 | 4 | 4 | 0 | 0 | 100.00% | 100.00% | Negative |
| CVS002 | 5 | 5 | 0 | 0 | 100.00% | 100.00% | Positive |
| CVS004 | 2 | 2 | 0 | 0 | 100.00% | 100.00% | Negative |
| CVS006 | 17 | 16 | 1 | 0 | 94.12% | 94.12% | Negative |
| CVS008 | 25 | 25 | 0 | 0 | 100.00% | 100.00% | Negative |
| CVS010 | 3 | 2 | 1 | 0 | 66.67% | 66.67% | Negative |
| CVS011 | 8 | 7 | 1 | 0 | 87.50% | 87.50% | Negative |
| CVS012 | 14 | 14 | 0 | 0 | 100.00% | 100.00% | Negative |
| CVS013 | 6 | 6 | 0 | 0 | 100.00% | 100.00% | Negative |
| CVS014 | 3 | 3 | 0 | 0 | 100.00% | 100.00% | Negative |
| CVS015 | 14 | 14 | 0 | 0 | 100.00% | 100.00% | Negative |
| CVS016 | 8 | 7 | 1 | 0 | 87.50% | 87.50% | Negative |
| CVS017 | 7 | 4 | 3 | 0 | 57.14% | 57.14% | Negative |
| CVS018 | 10 | 9 | 1 | 0 | 90.00% | 90.00% | Positive |
| CVS020 | 4 | 3 | 1 | 0 | 75.00% | 75.00% | Positive |
| CVS022 | 3 | 2 | 1 | 0 | 66.67% | 66.67% | Positive |
| CVS023 | 4 | 4 | 0 | 0 | 100.00% | 100.00% | Negative |
| CVS024 | 4 | 1 | 3 | 0 | 25.00% | 25.00% | Positive |
| CVS025 | 5 | 5 | 0 | 0 | 100.00% | 100.00% | Negative |
| CVS026 | 3 | 2 | 1 | 0 | 66.67% | 66.67% | Negative |
| CVS027 | 4 | 4 | 0 | 0 | 100.00% | 100.00% | Negative |
| CVS028 | 4 | 4 | 0 | 0 | 100.00% | 100.00% | Negative |
| CVS029 | 3 | 3 | 0 | 0 | 100.00% | 100.00% | Negative |
| CVS030 | 7 | 7 | 0 | 0 | 100.00% | 100.00% | Negative |
| CVS032 | 3 | 1 | 2 | 0 | 33.33% | 33.33% | Negative |
| CVS034 | 8 | 6 | 2 | 0 | 75.00% | 75.00% | Negative |
| CVS035 | 6 | 6 | 0 | 0 | 100.00% | 100.00% | Negative |
| CVS036 | 8 | 1 | 7 | 0 | 12.50% | 12.50% | Negative |
| CVS037 | 7 | 7 | 0 | 0 | 100.00% | 100.00% | Negative |
| CVS038 | 7 | 0 | 7 | 0 | 0.00% | 0.00% | Positive |
| CVS039 | 2 | 0 | 2 | 0 | 0.00% | 0.00% | Positive |
| CVS040 | 8 | 8 | 0 | 0 | 100.00% | 100.00% | Positive |
| CVS041 | 5 | 5 | 0 | 0 | 100.00% | 100.00% | Negative |
| CVS042 | 6 | 2 | 4 | 0 | 33.33% | 33.33% | Positive |
| CVS043 | 6 | 6 | 0 | 0 | 100.00% | 100.00% | Negative |
| CVS044 | 4 | 4 | 0 | 0 | 100.00% | 100.00% | Negative |
| CVS045 | 5 | 3 | 2 | 0 | 60.00% | 60.00% | Negative |
| CVS046 | 7 | 5 | 2 | 0 | 71.43% | 71.43% | Positive |
| CVS047 | 5 | 5 | 0 | 0 | 100.00% | 100.00% | Negative |
| CVS048 | 3 | 3 | 0 | 0 | 100.00% | 100.00% | Negative |
| CVS049 | 3 | 3 | 0 | 0 | 100.00% | 100.00% | Negative |
| CVS050 | 9 | 6 | 3 | 0 | 66.67% | 66.67% | Negative |
| CVS051 | 7 | 7 | 0 | 0 | 100.00% | 100.00% | Negative |
| CVS052 | 7 | 4 | 3 | 0 | 57.14% | 57.14% | Negative |
| CVS053 | 8 | 8 | 0 | 0 | 100.00% | 100.00% | Negative |
| Total | 291 | 243 | 48 | 0 |  |  |  |

**Table 5.** Patient details and classification for LD-REIMS data

| Patient | Number of spectra | Number of passes | Number of failures | Number of outliers | Correct Classification Rate (Exluding outliers) | Correct Classification Rate (Including outliers) | True Classification |
| --- | --- | --- | --- | --- | --- | --- | --- |
| CVS003 | 5 | 2 | 3 | 0 | 40.00% | 40.00% | Positive |
| CVS006 | 18 | 18 | 0 | 0 | 100.00% | 100.00% | Negative |
| CVS007 | 3 | 1 | 2 | 0 | 33.33% | 33.33% | Positive |
| CVS008 | 12 | 9 | 3 | 0 | 75.00% | 75.00% | Negative |
| CVS009 | 7 | 7 | 0 | 0 | 100.00% | 100.00% | Positive |
| CVS010 | 6 | 4 | 2 | 0 | 66.67% | 66.67% | Negative |
| CVS011 | 6 | 0 | 6 | 0 | 0.00% | 0.00% | Negative |
| CVS012 | 11 | 9 | 2 | 0 | 81.82% | 81.82% | Negative |
| CVS014 | 7 | 7 | 0 | 0 | 100.00% | 100.00% | Negative |
| CVS016 | 2 | 2 | 0 | 0 | 100.00% | 100.00% | Negative |
| CVS017 | 6 | 6 | 0 | 0 | 100.00% | 100.00% | Negative |
| CVS018 | 6 | 0 | 6 | 0 | 0.00% | 0.00% | Positive |
| CVS019 | 7 | 7 | 0 | 0 | 100.00% | 100.00% | Positive |
| CVS020 | 7 | 6 | 1 | 0 | 85.71% | 85.71% | Positive |
| CVS021 | 3 | 3 | 0 | 0 | 100.00% | 100.00% | Positive |
| CVS022 | 3 | 3 | 0 | 0 | 100.00% | 100.00% | Positive |
| CVS023 | 2 | 2 | 0 | 0 | 100.00% | 100.00% | Negative |
| CVS024 | 3 | 3 | 0 | 0 | 100.00% | 100.00% | Positive |
| CVS048 | 5 | 4 | 1 | 0 | 80.00% | 80.00% | Negative |
| CVS049 | 3 | 3 | 0 | 0 | 100.00% | 100.00% | Negative |
| CVS050 | 5 | 5 | 0 | 0 | 100.00% | 100.00% | Negative |
| CVS051 | 6 | 6 | 0 | 0 | 100.00% | 100.00% | Negative |
| CVS052 | 4 | 4 | 0 | 0 | 100.00% | 100.00% | Negative |
| CVS053 | 8 | 7 | 1 | 0 | 87.50% | 87.50% | Negative |
| CVS055 | 2 | 0 | 2 | 0 | 0.00% | 0.00% | Positive |
| Total | 147 | 118 | 29 | 0 |  |  |  |
